# Cone Rescue with Laser Photobiomodulation in Murine and Human Retinal Dystrophy

**DOI:** 10.1101/2021.04.08.21255175

**Authors:** Robert J. Casson, John P.M. Wood, Jack Ao, Jagjit S. Gilhotra, Shane R. Durkin, WengOnn Chan, Glyn Chidlow

## Abstract

Retinitis pigmentosa (RP) encompasses a genetically diverse group of blinding inherited retinal diseases. In most subtypes the gene defect is expressed in the rod photoreceptors, yet in many affected individuals the cone photoreceptors undergo secondary degeneration, leading to loss of the remaining central vision. There is evidence that bioenergetic and oxidative stress are involved in this secondary cone loss. Photobiomodulation (PBM) uses low energy light in the far red or near-infrared spectrum to manipulate cellular activity. We have used a novel slit lamp-mounted retinal PBM laser to deliver precise energy levels to targeted retina. We showed that PBM laser attenuates oxidative and bioenergetic stress-induced photoreceptor loss *in vitro* and rescues cones in the *rd1* murine model of RP. In a phase I trial (ACTRN12618000651280), foveal laser treatment was safe in humans with RP and temporarily recovered, on average, 5 letters of visual acuity.

## Introduction

Retinitis pigmentosa (RP) refers to a genetically heterogenous group of inherited retinal dystrophies (IRDs) united by a characteristic clinical phenotype. In most affected individuals, the cone photoreceptors do not express the genetic defect, and yet, in most of these individuals, secondary cone degeneration ensues, resulting in loss of central vision and profound visual impairment. There is no clinically proven therapy that preserves vision in RP and the IRDs have overtaken diabetic retinopathy as the leading cause of blindness in the working age population in certain regions. 1 Bioenergetic dysfunction and oxidative stress are strongly implicated in the pathogenesis of cone degeneration in RP; 2, 3, 4 hence, cone mitochondria are an attractive therapeutic target. An approach that simultaneously enhances oxidative energy metabolism and reduces reactive oxygen species (ROS) production in cones is particularly appealing and has the added benefit that it is not contingent upon the specific rod gene defect, with potentially broad clinical applicability. 4

Photobiomodulation (PBM) refers to the treatment of tissue with light in the far red to near-infrared spectrum (630-1000 nm). Most evidence supports an action of PBM on mitochondrial metabolism, particularly on cytochrome *c* oxidase in the electron transport chain. 5, 6, 7 An increasingly substantive body of work has revealed positive effects of PBM on photoreceptor survival. 8 To date, studies have almost invariably used light emitting diode (LED) systems; however, this methodology suffers the serious disadvantage that the energy reaching the retina is uncontrolled. In the current study, we utilized an experimental 670 nm slit lamp-delivered retinal laser that enables controlled delivery of a known intensity (irradiance) and energy density (fluence) to the retina. Here, we report the effect of this novel laser on photoreceptor preservation in retinal cultures and on cone survival in a murine model of severe RP. In addition, we translated this technology to a phase I study in human subjects with RP and we herein report the results.

## Results

### Preservation of rod and cone photoreceptors in culture by PBM

We utilised retinal culture preparations which have previously been shown to contain a wide variety of cell populations including inner retinal neurons, rhodopsin-expressing neurons (designated as rods) and S opsin-expressing neurons (representing S-cones). 9, 10 We assessed the effect of PBM on cultures under conditions of oxidative stress (via application of the peroxide analogue, tert-butyl hydroperoxide; tbH) and mitochondrial compromise (via the complex IV inhibitor, sodium azide). Treatment of cells with PBM alone for 90 seconds did not detrimentally affect neurons or photoreceptors at exposures up to 100 mW/cm^2^. When elevated to 250 mW/cm^2^, however, there was a tendency for reduction in both rod and S-cones. In the case of the S-cones, this cell loss became statistically significant at 450 mW/cm^2^ (Supplemental Figure 1). For this reason, cell culture studies were conducted using a maximum PBM exposure of 100 mW/cm^2^.

Exposure of cultures to oxidative stress or mitochondrial compromise for 24 h resulted in dramatic reduction in rod cell labelling (Figure 1A). Pre-treatment with PBM, 6 hours prior to addition of toxins, at 100 mW/cm^2^ but not at 25 mW/cm^2^ significantly attenuated cell loss from both oxidative stress and mitochondrial dysfunction (Figure 1B). Similar results were found with S-opsin-labelled cones (Figure 1C). Again, pre-treatment with PBM laser significantly prevented cell loss when applied at 100 mW/cm^2^ but not at 25 mW/cm^2^ (Figure 1D). These data demonstrate the protective effect of PBM on compromised retinal cells and imply that this effect is afforded by attenuation of oxidative stress and mitochondrial dysfunction.

**Figure 1.**
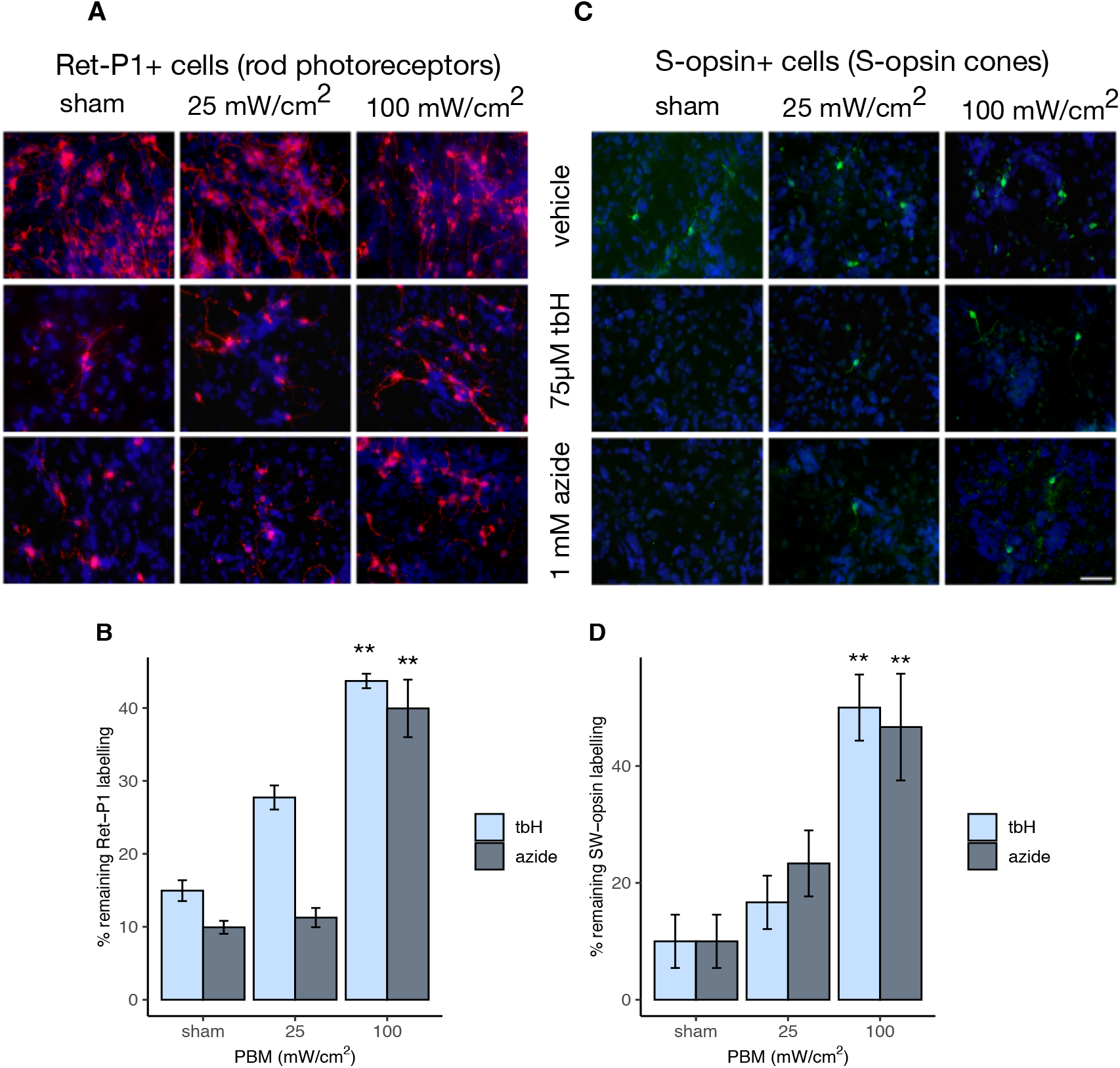
Protection of photoreceptor cells in mixed retinal cultures by PBM. Mixed cultures of retinal cells, consisting of neurons, glia and photoreceptors, were exposed *in vitro* to PBM at either 25 or 100 mW/cm^2^, or to sham treatment. After 6 hours, they were subjected to either oxidative stress (75 µM tert-butyl-hydroperoxide; tbH) or mitochondrial compromise (1 mM sodium azide) for a further 24 hours. Immunocytochemical labelling for rod photoreceptors (A, B) and for S-cones (C, D) was significantly decreased by exposure of cultures to both tbH and azide. Pre-treatment with PBM at 100 mW/cm^2^, but not at 25 mW/cm^2^, significantly alleviated the effects of both toxins for each cell-type compared to sham. ^*^*P* < 0.05, ^**^*P* < 0.01, by post hoc Dunnett’s test compared to sham group; n = 6 determinations for each test group; error bars depict SEM. Scale bar: 50 µm.

Given the potential pathogenic role of oxidative stress and bioenergetic compromise in RP 2, 3, 4 and the potential translatability of PBM-induced cone rescue in human RP, we then progressed to *in vivo* investigation of the effects of PBM on retinal cones in *rd1* mice.

### Preservation of M/L- and S-cones in rd1 mice by PBM

We initially assessed the effect of PBM on cone rescue at P60, using cone cell immunostaining density averaged across each retinal flatmount as the primary outcome. Mice received twice weekly PBM treatment to one eye commencing at P21. At P60, M/L-opsin^+^ cone density was significantly greater in *rd1* mice treated with PBM at either 25 mW/cm^2^ or 100 mW/cm^2^ compared to shams (Figure 2A, Supplemental Figure 2). S-opsin^+^ cones were similarly rescued (Figure 2B, Supplemental Figure 2). These data indicate that cone survival was prolonged by PBM, but do not shed light on whether cones retain any functionality. Hence, we also assessed survival of cone outer segments, whose presence is required for detecting light. In *rd1* mice, cone segments are typically immunolabeled with greater fluorescent intensity than cell bodies, which permits separate quantification of segments from cell bodies by image thresholding (Supplemental Figure 2), and they degenerate far more rapidly than their somas. The data showed that cone outer segment survival was likewise significantly prolonged by PBM (Figure 2C). Electroretinographic activity in the *rd1* mouse is unrecordable by P30 due to the early onset of cone segment degeneration. 11 Hence, as it was deemed unethical to subject pups to multiple episodes of general anesthesia and PBM treatments, we assessed residual vision by recording the optokinetic (OKN) reflex at P35 and observed partial preservation of this visual reflex in the PBM treated groups (Figure 2D). Of note, there was no significant difference in M/L-opsin or S-opsin cone density or OKN preservation between mice treated with 25 mW/cm^2^ and those treated with 100 mW/cm^2^ PBM (Figure 2A-D).

**Figure 2.**
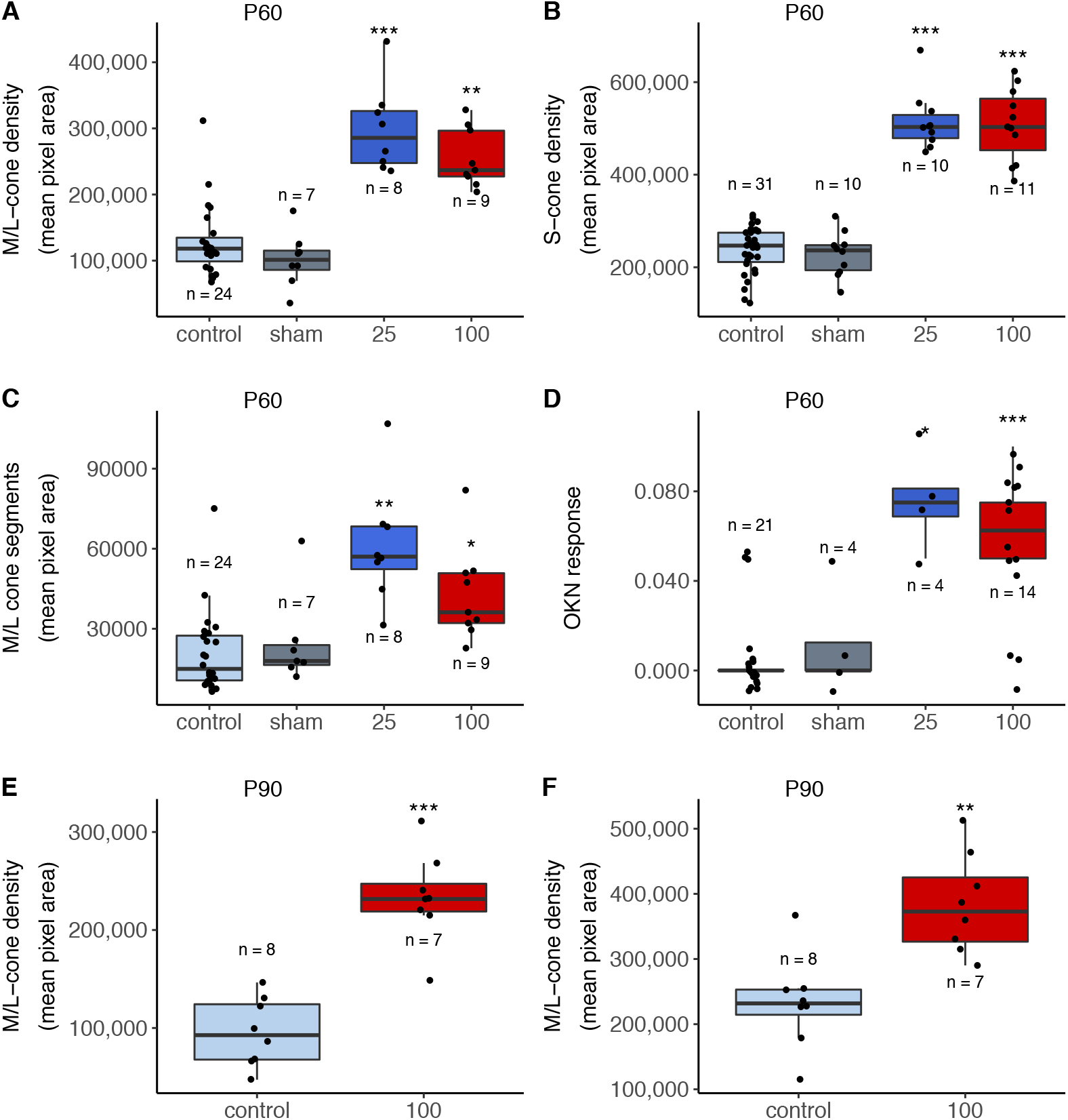
Cone rescue by PBM in *rd1* mice at P60 and P90. M/L-opsin^+^ (**A**) and S-opsin^+^ (**B**) cone densities of *rd1* mice treated with PBM at irradiances of either 25 mW/cm^2^ or 100 mW/cm^2^ were significantly enhanced compared to untreated control and sham-treated mice. M/L-opsin^+^ cone outer segments were also significantly preserved (**C**). The optokinetic reflex was significantly preserved, indicating functional preservation of vision in *rd1* mice treated with PBM (**D**). Both M/L-opsin^+^ and S-opsin^+^ cones were also rescued at P90 (**E, F**). Box length = interquartile range (IQR). Black horizontal line = median, whiskers = 1.5 x IQR; black circles = data points. *** p < 0.001; ** p < 0.01, * p < 0.05, by post hoc Dunnett’s test compared to sham/control group.

To determine whether the neuroprotective influence of PBM extended to longer durations, we then investigated cone survival at P90, with mice again receiving twice weekly PBM treatment to one eye. Given that there was no significant irradiance level difference in efficacy at P60, we elected to treat solely with 100 mW/cm^2^. We again observed marked preservation of cones compared to untreated controls (Figure 2E, 2F).

Having noted the remarkable protective effect on *rd1* cones and lack of toxicity from our previous study, 12 we rapidly translated this technology to a phase I trial of patients with advanced RP that had progressed to tunnel vision and impairment of cone-derived visual acuity.

### Phase I study

A total of 25 patients from RJCs, JSGs and SRDs private clinical practices and the Retinal clinics at the Royal Adelaide Hospital (RAH) were identified as potential participants; 12 declined to participate and 13 were screened. One patient failed screening because the visual acuity was still within normal limits (20/30 in the worst eye); 12 patients were entered into the study and designated as Group 1 or Group 2 according to enrolment chronology (Figure 3).

**Figure 3.**
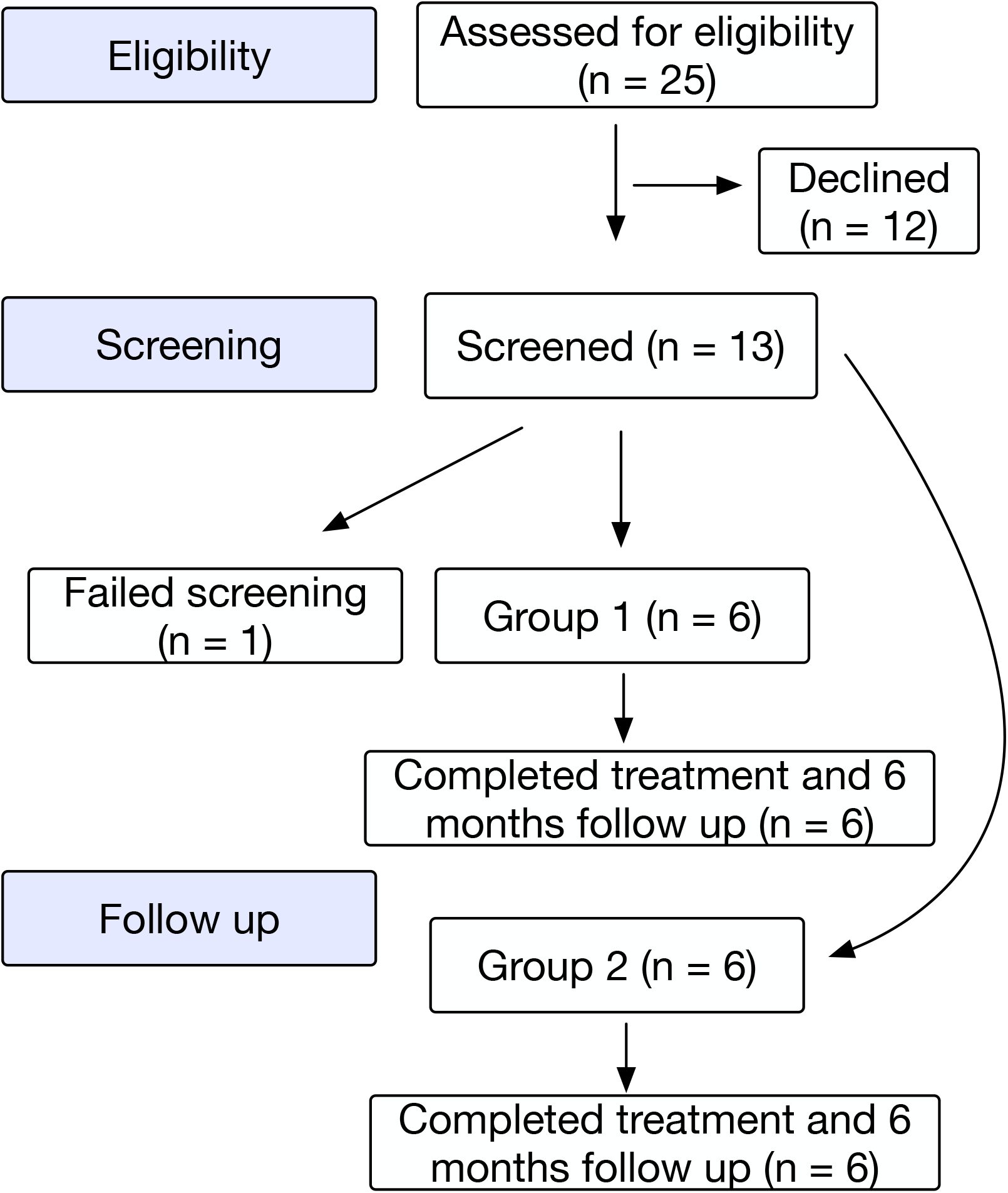
Participant flow chart.

The procedure was extremely well tolerated by all participants and there were no adverse events at either the 25 mW/cm^2^ or the 100 mW/cm^2^ settings. There were no missing data. Combining both groups, patients recovered a mean of 5.4 letters (SD 5.5) at the 8-week time point (4 weeks after completion of treatment, Figure 4A). The group receiving 100 mW/cm^2^ displayed a greater variance (Figure 4C). Cone-derived photopic flicker responses were almost completely abolished in all participants with a large within-subject and between-subject variance (Figure 4B). No significant change in ERG amplitude was observed (Figure 4D, F). The procedure had no significant effect on any of the measured ophthalmic parameters apart from visual acuity. There was a suggestion that the effect on visual acuity was tapering by 6 months (Figure 4E)

**Figure 4.**
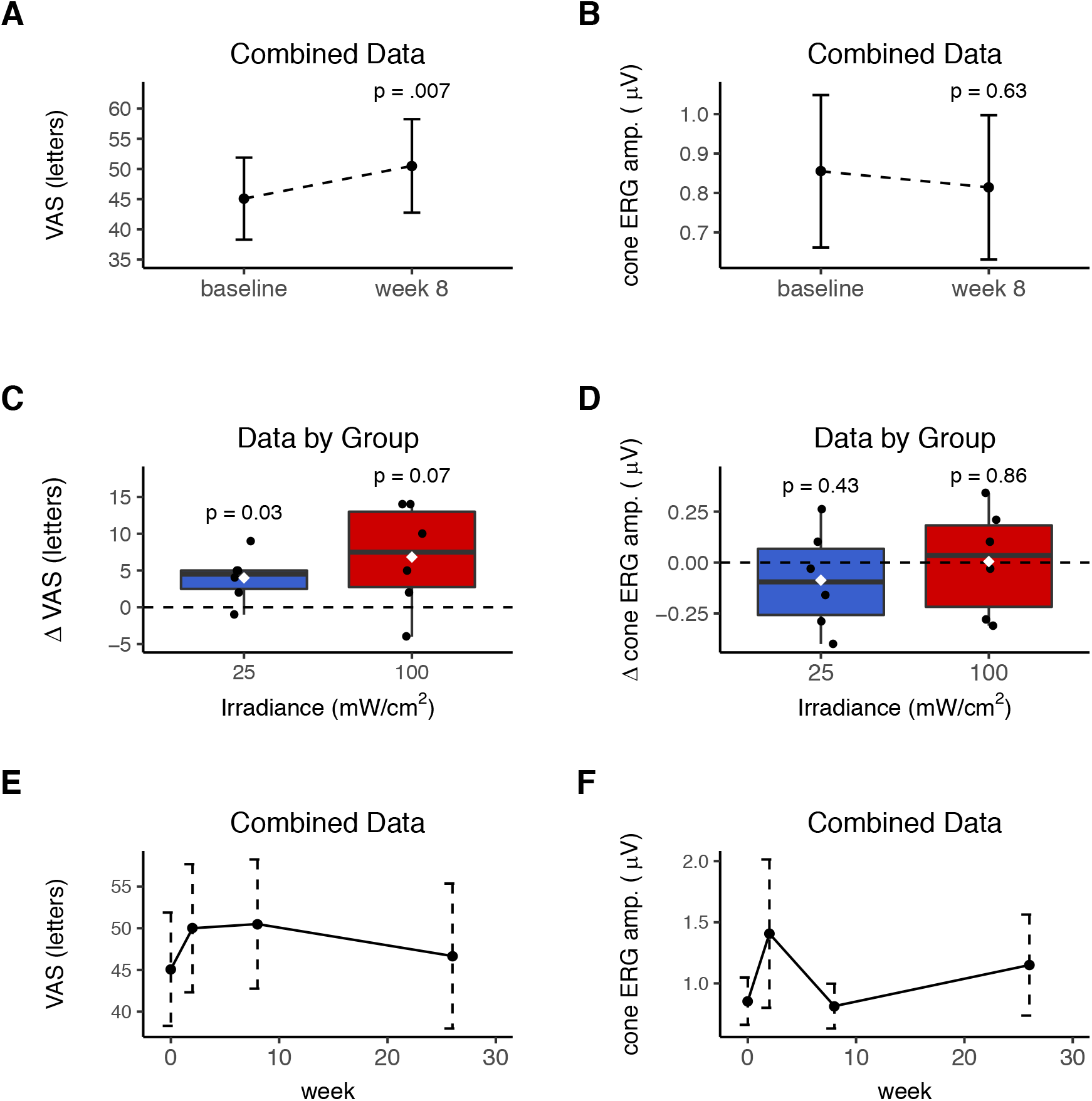
Effect of PBM on visual acuity and electroretinographic (ERG) photopic flicker amplitude (amp.) in individuals with RP. (**A**) Combining data from both groups, PBM significantly recovered the visual acuity score (VAS) by a mean of 5 letters at the pre-defined time point, 4 weeks after the final treatment (week 8; *n*= 12). (**C-D)** VAS and ERG data by irradiance group (*n* = 6 per group with p values comparing week 8 to baseline). (**E-F**) Combined data (*n* = 12) showing change in VAS and the photopic flicker amp. from baseline to the 26-week endpoint. Error bars depict standard errors. Box length = interquartile range (IQR). Black horizontal line = median, white diamond = mean, whiskers = 1.5 x IQR; black circles = individual data points. P values calculated using Welch’s *t*-test (VAS comparisons) or Wilcoxon test (ERG data) on paired data.

## Discussion

PBM invokes an array of tissue responses that make it an attractive experimental therapeutic modality for retinal diseases that comprise bioenergetic failure, oxidative stress and/or gliosis components. 13 PBM from 670 nm LED devices has been shown to mitigate photoreceptor injury in a range of animal models, including methanol toxicity, 14 and oxygen-induced degeneration. 15, 16 A recent study demonstrated the safety and feasibility of PBM as a treatment for retinopathy of prematurity in neonatal intensive care. 17 Retinal protection against white light-induced degeneration has been reported, 18, 19 including reduced glial 20 and complement activation 21 and specific cone protection. 22 In 2013, Kirk *et al*. reported that 670 nm LED light (Quantum Devices, Barneveld, WI) attenuated loss of retinal function and structure in a rat model of RP (the P23H rat). 23 More recently, this team demonstrated the protective effect of 830 nm LED light on the retina of the P23H rats. 24 These authors demonstrated that PBM preserved the mitochondrial redox state (NADH/FAD ratio) and provided partial protection against electroretinographic dysfunction and histological loss of photoreceptors. 24 Although cones were not specifically investigated in these studies, the results are consistent with the findings from the current study.

The *rd*^*Pde6b-/-*^ *(rd1)* mouse is a well-characterized model of autosomal recessive RP that carries a murine leukemia virus (Xmv-28) insert in intron 1 and point-nonsense mutation in exon 7, eliminating the catalytic domain of the phosphodiesterase protein product (PDE6B). 25 In the *rd1* retina, this mutation causes the rapid degeneration of rods, commencing at P8-10 26 and complete by P40. 27 Cones do not express *PDE6B*; however, a chronic, secondary degeneration of cones ensues, mirroring the human phenotype. 9

Most non-primate mammals, including mice, have two types of cone opsin with distinct spectral absorbance profiles: short-wavelength sensitive (S) opsin and medium/long-wavelength (M/L) sensitive opsin. In wild type mice, the S- and M/L-opsins are expressed in the outer segments, and many cones in the mouse retina express both opsins (dual cones). 28 We and others 9, 29 have noted that cone outer segment (OS) loss is an early feature of cone degeneration in *rd1* mice and is accompanied by ectopic redistribution of opsins to the remaining cell soma. Hence, immunolocalization of opsin must be interpreted in this histomorphological context. We have also previously reported that the loss of both S- and M/L-OS is complete by P21 in the central retina, and 90% complete by P45 in the peripheral retina. 9

The IRDs remain a significant visual health problem at the individual and societal level and have traditionally been recalcitrant to therapeutic intervention. However, recent technological advances have made some progress. Based on the results of improved multi-luminance mobility testing in a phase III randomized controlled trial (RCT), 30 the Food and Drug Administration recently approved voretigene neparvovec-rzyl (LUXTURNA®; Spark Therapeutics, Philadelphia, PA, USA) for the treatment of patients with confirmed biallelic *RPE65-*mediated IRD. Voretigene is administered via subretinal injection requiring a pars plana vitrectomy with a current cost of $425,000 per eye. 31 RPE65 mutations account for approximately 0.1-1% of IRD. Gene therapy for other recessive IRDs is an explosive research area. Similarly, sophisticated genetic techniques utilising the CRISPR/Cas 9 system to manipulate toxic gain-of-function autosomal dominant mutations are undergoing investigation. 32 However, these genetic engineering techniques are disease specific, likely to be extremely expensive, and restricted to high socio-economic index populations. The development of artificial retinal implants is largely targeted at IRDs but has had limited clinical impact to date. Less expensive universal strategies, including dietary supplements of vitamin A and fish oils have been trialled in RCTs, but have produced limited changes in the natural history of RP and inconsistent results. 33 Recently, Campochiaro *et al*. reported that oral N-acetyl-cysteine (NAC) improved cone function in a phase I trial on individuals with RP. 34 The mean BCVA improved 2-3 letters in each of the 3 cohorts of 10 patients receiving different doses of NAC over a 24-week period. 34 Hence, the 5-letter short-term improvement noted in the current study compares favourably with oral anti-oxidants.

A relatively low-cost strategy that preserves central vision irrespective of the rod gene defect would be a major medical breakthrough at the individual and community level. We have recently demonstrated that retinal PBM laser reduces diabetic macular edema (DME) 12 and the Diabetic Retinopathy Clinical Research Network (DRCR.net) is currently conducting a National Eye Institute-sponsored larger scale RCT of LED-delivered PBM versus sham in DME. Retinal PBM is an intriguing therapeutic strategy and the results from the current translational research strongly motivate further studies investigating PBM as a treatment for RP. In particular, further work is required to determine the optimal frequency of delivery and to assess the effect in a suitably powered RCT.

## Methods

Detailed laboratory methods are described in Supplemental Methods.

### Clinical Trial Primary Outcome Measure

We conducted a phase 1 single ascending dose study (ACTRN12618000651280), assessing the safety of PBM to the macula in individuals with RP. The Transparent Reporting of Evaluations with Nonrandomized Designs (TREND) guidelines were followed. 35 The primary outcome measure was the safety profile, and the secondary outcomes were the change in visual acuity and photopic flicker electroretinogram (ERG). The target population was individuals with advanced RP and partial loss of central vision.

### Participants and Protocol

We aimed to include a total of 12 patients in the clinical trial. Our protocol designated 6 patients to receive the lower PBM irradiance setting (25 mW/cm^2^; Group 1) and, upon completion of the treatment and safety determined after 6 months follow-up, a further 6 patients would be designated to the higher PBM irradiance setting (100 mW/cm^2^; Group 2).

Irradiance parameters were selected based on our previous animal safety studies and on settings used in our previous clinical trial assessing the effect of laser PBM on DME. 12

The data collection and laser treatment were conducted in the Ophthalmology Department at the RAH. Patients had to be ≥ 18 years of age and have a best-corrected visual acuity (BCVA) between 20/40 and 20/800 (5 – 70 letters) with visual impairment attributed to RP. Patients were excluded if they had had an intraocular procedure or laser treatment within the previous 6 months, or if they had cystoid macula edema. Patients underwent a baseline assessment (week 0) then received 2 treatments per week (separated by approximately 48 hours) for 4 weeks. Patients were encouraged to attend all visits and were given verbal and written reminders of ongoing appointments. At baseline, the best-corrected logarithm of the minimum angle of resolution (logMAR) visual acuity was recorded using an Early Treatment Diabetic Retinopathy (ETDRS) chart at 4 m (or 1 m if the BCVA was < 20/200) Slit lamp examination of the anterior segment was performed by an experience ophthalmologist (RJC) and intraocular pressure measured with Goldman tonometry. The pupil was dilated with tropicamide 0.5% and the fundus examined. Spectral domain optical coherence tomography (SD-OCT) of the macula using the Cirrus™ HD-OCT (Carl Zeiss Meditec AG, Dublin CA), fundus photography and photopic flicker ERG with the RETeval^®^ system (LKC Technologies, Gaithersburg, MD) were performed. Baseline assessments were repeated after 2, 8 and 24 weeks. The visual acuity was measured by a single experienced ophthalmic nurse and the patient was encouraged to read as many letters as possible at each visit. As per the ETDRS chart protocol, the examiner stopped the test only when it became evident that no further meaningful readings were being made, despite urging the subject to guess.

The development of any ocular adverse reaction and/or reduction in visual acuity of ≥ 5 letters at the 2-week visit mandated withdrawal. Any ocular adverse effects in Group I prohibited proceeding to Group II. Any undesirable clinical occurrence in a patient whether it was considered to be device related or not, that included a clinical sign, symptom or condition and/or an observation of an unintended technical performance or performance outcome of the device were carefully recorded.

### Treatment

The pupil was dilated with tropicamide 0.5% and the cornea anesthetized with topical amethocaine. At the slit lamp, a Mainster focal/grid lens was used to visualize the fundus. Each treatment consisted of a 90 s exposure of the macular region to the 4.5 mm diameter laser beam. The laser intensity was set at 25mW/cm^2^ for the low dose (Group I) patients and 100 mW/cm^2^ for high dose (Group II) patients.

### Study approval

The experiments involving animals adhered to the Australian code for the care and use of animals for scientific purposes (the Code) and were approved by the University of Adelaide Animal Ethics Committee. The clinical study was conducted in accordance with the Declaration of Helsinki and was approved by the Central Adelaide Local Health Network Human Research Ethics Committee. Written informed consent was received from the participants prior to inclusion in the study.

### Statistics

Initial exploratory analyses were performed on all data including construction of box plots and examination of the distribution and heterogeneity of residuals with histograms and Q-Q plots. Where normality was met, comparisons between groups were made with either a post hoc Dunnett’s test or Welch’s paired *t*-test. If normality was not met a non-parametric test was used. A p value < 0.05 was considered statistically significant. Statistical analyses were performed using the R statistical software. 36

## Data Availability

The datasets generated during and/or analysed during the current study are available from the corresponding author on reasonable request.

## Author contributions

GC, JPMW and JA conceived and performed the laboratory-based experiments. RJC, GC, JPMW and JA performed the laboratory-based data analyses. RJC, JSG, SRD and WC conducted the clinical trial. RJC performed the clinical trial analyses and led the writing of the manuscript. All authors contributed to drafting of the manuscript.

## Acknowledgements

This research was sponsored in part by a National Health & Medical Research Council grant: (APP1102568) “Novel Photoreceptor Bioenergetics: Basic Science & Clinical Translation” and a grant from the Ophthalmic Research Institute of Australia (ORIA): “Rescuing Cone Photoreceptors in Retinitis Pigmentosa with Photobiomodulation”.

