## Supplementary Material for "Cone Rescue with Laser Photobiomodulation in Murine and Human Retinal Dystrophy"

**Supplementary Methods**

*Photobiomodulation Retinal Laser*

The photobiomodulation (PBM) laser system comprised a custom-designed, slit lamp microscope-mounted Integre near-infrared laser (Ellex Medical Lasers, Adelaide, South Australia, Australia) incorporating a 670-nm light source emitting a beam 4.5 mm in diameter with a flat-top profile.

*Cell Culture Studies*

Cultures were prepared by enzymatic and mechanical dissociation of newborn Sprague-Dawley rat pups, as previously described. (1) Cultures comprised the majority of cell-types present in the newborn rat retina, including neurons, short wave opsin-expressing cones (S-cones) and rhodopsin-labelling cells, designated as rods. Cells were seeded onto borosilicate glass coverslips pre-coated with 10 μg/ml poly-L-lysine and maintained in standard Minimal Essential Medium (MEM) containing phenol red indicator, supplemented with 10 % (v/v) fetal bovine serum and antibiotics for 7-8 days.

For laser treatment, coverslips with adherent cells were placed into culture plates containing serum-free MEM lacking phenol red, and positioned on a pre-warmed, bespoke, horizontal platform attached to the chin-rest of the slit lamp delivery system. Treatments were applied centrally to individual coverslips at a range of radiant exposures (25, 100, 250, 450 $mW/cm^2$) for 90s. The area covered by PBM treatment was of 15.9 $mm^2$ (4.5 mm diameter). Sham treatment was carried out in the same manner as for PBM application except that exposure was only to the laser aiming beam for the allotted time and not the laser. After treatment, cells were replaced into standard medium.

In some experiments, 24 h after PBM exposure, one of two stressors was added: 75 μM tert-butyl hydroperoxide (tbH), to induce oxidative stress, or 1 mM sodium azide, to compromise mitochondrial function. Stressors were left for a further 24 hours before cultures were fixed with neutral-buffered formalin (15 minutes) and processed for immunocytochemistry as described previously. (1) Cultures were immunocytochemically labelled as follows: neurons for tau (Dako, Denmark; 1:5000), rod photoreceptors for rhodopsin (clone Ret-P1; Santa Cruz Biotechnology In., Santa Cruz, USA; 1:5000) and SW cones for S-opsin (sc-14363, Santa Cruz Biotechnology Inc., Santa Cruz, USA; 1:1000). Nuclear counter-staining of cells was achieved with a final five-minute incubation of coverslips in 500 ng/ml 4′,6-diamidino-2-phenylindole (DAPI). Images were collected from each of 6-8 different cultures for each treatment and Image-J software (NIH, Bethesda, Maryland, USA) subsequently used to quantify labelling for each specific cell-type being investigated. Data were compared using one-way analysis of variance followed by Tukey's test for multiple comparisons; significance was denoted by $P < 0.05$.

*Animal Studies*

Mice were divided into three treatment groups: sham (aiming beam only), lower dose PBM laser (25 $mW/cm^2$) and higher dose PBM laser (100 $mW/cm^2$). Since the photoreceptor loss occurs bilaterally, one eye received PBM laser treatment while the fellow untouched eye served as a paired control. Treatment began from P21 and occurred twice weekly until euthanasia. Topical oxybuprocaine was applied to the ocular surface and the pupil dilated with tropicamide. Mice received inhalational anesthesia with isoflurane delivered via a nosecone throughout the procedure. A coverslip was placed on the cornea and mice were positioned on a custom-designed platform at the slit lamp laser delivery system. The laser was centered on the optic nerve and the fundus was exposed to PBM laser for 90 s.

*Optokinetic (OKN) Response*

The OKN reflex is commonly used to measure the visual function of an animal by observing the maximum spatial frequency of a rotating visual stimulus for which the animal responds with a head reflex. Animals were placed on a platform on a floor mirror surrounded by computer monitors that formed an enclosed area. Vertical sine wave gratings (100% contrast) were projected on the computer monitors. The spatial frequencies tested were 0.05, 0.075, 0.1, 0.2, 0.3, 0.4, 0.5, and 0.6 cycles per degree (cpd). A camera was placed above the platform to observe and record the animal's head movements. Mice were placed one at a time on the platform and allowed to acclimatize to their surroundings before starting the stimulus. The stimulus consisted of a grating perceptible to the mouse that was projected on the cylinder wall which rotated at a constant 12 degrees /second. Two independent experimenters masked to the treatment groups monitored the head reflex characterized by the animal displaying reflexive head movements corresponding to the direction and speed of the cylinder rotation that was not accompanied by any other body movements. A positive response was recorded only if the reflexive head movement occurred within the first 15 seconds of the stimulus and there was agreement between the experimenters. Assessment of the left or right eye was dependent on cylinder direction. Clockwise direction of the cylinder corresponded to the left eye whilst counterclockwise direction was related to the right eye. The spatial frequency of the grating was progressively increased until the animal no longer responded. The maximum spatial frequency for a positive head reflex was recorded. This was repeated for each eye.

*Tissue processing and immunohistochemistry*

All mice were euthanized by transcardial perfusion with physiological saline under terminal anaesthesia (100 mg/kg body weight ketamine and 10 mg/kg body weight xylazine) followed by decapitation. The superior aspect of each cornea was marked before globes were enucleated. For wholemount double labeling immunohistochemistry, eyes were fixed in 4% (w/v) neutral buffered formalin for 24 h and dissected into posterior eye-cups. The corneal mark was used to orient the eye and a small radial cut was made in the superior retina while it was still attached to the retinal pigment epithelium (RPE) in the eye-cup. Retinas were removed and prepared as flattened wholemounts by making another four radial cuts. Retinal wholemounts were then stored in phosphate buffered saline (PBS), prior to incubating in PBS containing 1% (v/v) Triton X-100 detergent (T) for 1 h at room temperature. Next, retinas were incubated in PBS-T containing 3% (v/v) normal horse serum (NHS-T) for 1 h at room temperature to block non-specific antibody binding. Retinas were then incubated overnight at 4 °C with a combination of primary antibodies diluted in NHS-T. OPN1SW antibody (1:1500, sc-14363, Santa-Cruz) was used to detect S-cones, whilst anti-R/G opsin antibody (1:1500, AB5405, Merck-Millipore) was used to detect M/L- cones. On day 2, retinas were washed for 1 h at room temperature in PBS-T, then incubated overnight at 4 °C with a combination of AlexaFluor-488 and -594 conjugated secondary antibodies (1:250; Invitrogen, Carlsbad, CA, USA) diluted in NHS-T. Finally, retinas were washed in PBS for 1 h at room temperature prior to mounting with the photoreceptor side facing up, using anti-fade mounting medium (Dako, Santa Clara, CA, USA).

*Image acquisition and quantification*

All analyses were conducted in a blinded fashion. Photomicrographs of wholemounts were taken with an epifluorescent microscope with attached fluorescent optics (BX-61; Olympus,

Mount Waverly, VIC, Australia). Rectangular areas of 526.5 x 422.5 μm were photographed adjacent to the optic disk and 2 mm away from the optic disk in each of the retinal quadrants. This yielded 8 images per retina (4 central and 4 peripheral). Quantification of cone survival was performed using Image-J software (NIH, Bethesda, Maryland, USA). Initially, however, images were processed in Photoshop CS3 (Adobe). Images were corrected for uneven lighting using a flatten filter and where necessary linear gradient tool, then sharpened, levels enhanced, and finally converted to 8-bit mode.

Cone cell bodies were identified by both their wide ovoid morphology, segments by their narrow, bundle-like appearance. For determination of total S-cone and total M/L-cone labelling, images were manually thresholded until all cone cell bodies and segments were highlighted. The area in pixels was then calculated with the "analyze particles" function, using a minimum size of 5 square pixels. As shown in supplemental Figure 3, ML-opsin segments stained with greater fluorescent intensity then cell bodies. This permitted quantification of M/L-opsin-labelled cone segment survival (which necessarily comprises both genuine M/L-cones and dual cones). Quantification of segments was performed by adjusting the image threshold to isolate the more intensely stained M/L-opsin$^{+}$ segments. The area in pixels was then calculated with the "analyse particles" function, using a minimum size of 5 square pixels. In contrast to M/L-opsin, S-opsin$^{+}$ cone cell bodies and segments stained with similar intensity. Separate quantification of S-opsin$^{+}$ segments from cell bodies could not be accurately attained with image threshold adjustment.

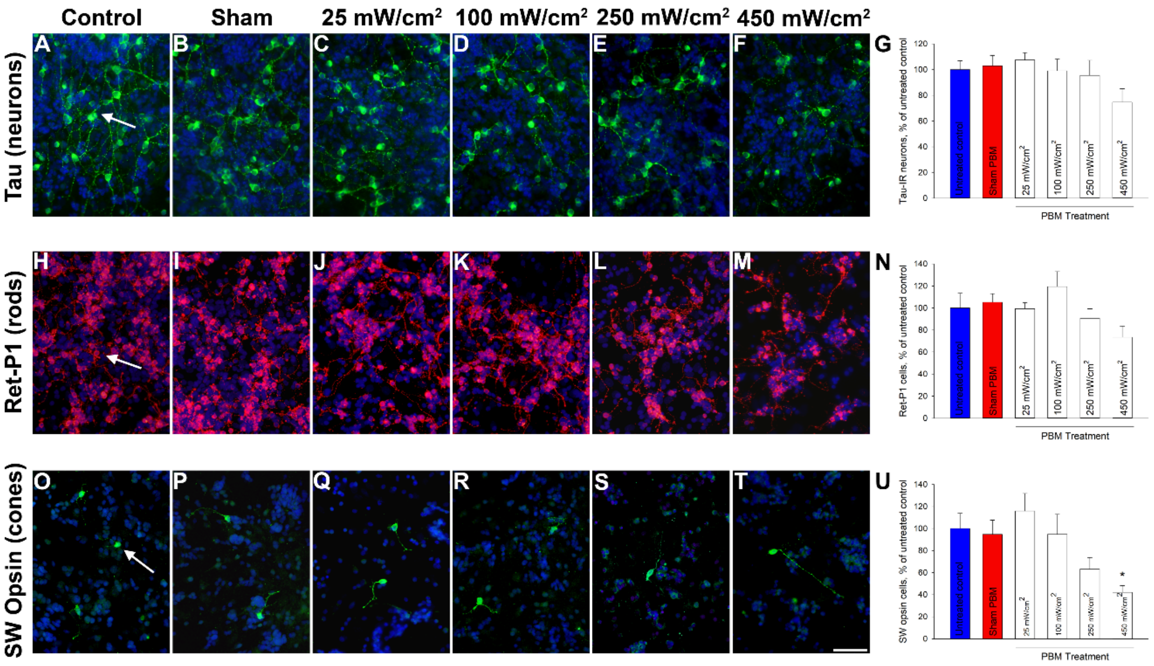


Supplementary Figure 1. Dosimetry response of retinal neurons and photoreceptors in culture to PBM. Cultures were exposed to 90 seconds of PBM laser in vitro at a range of energy settings and then fixed after 24 hours to determine potential cytotoxicity. Neither tau-labelled neurons (A-G) nor rhodopsin-labelled rod photoreceptors (H-N) were detrimentally affected by PBM laser application up to 250 mW/cm$^2$. At 450 mW/cm$^2$, however, there was a tendency for a reduction in labelling for both cell-types, although this effect was not of statistical significance. In the case of SW opsin-labelled cone photoreceptors (O-U), however, there was an apparent reduction when PBM was applied at above 100 mW/cm$^2$: this effect was determined to be significant at 450 mW/cm$^2$. $^*P < 0.05$ by one-way ANOVA plus Dunnett's test for multiple comparisons; $n$=6 determinations for each test group. Scale bar: 50 μm.

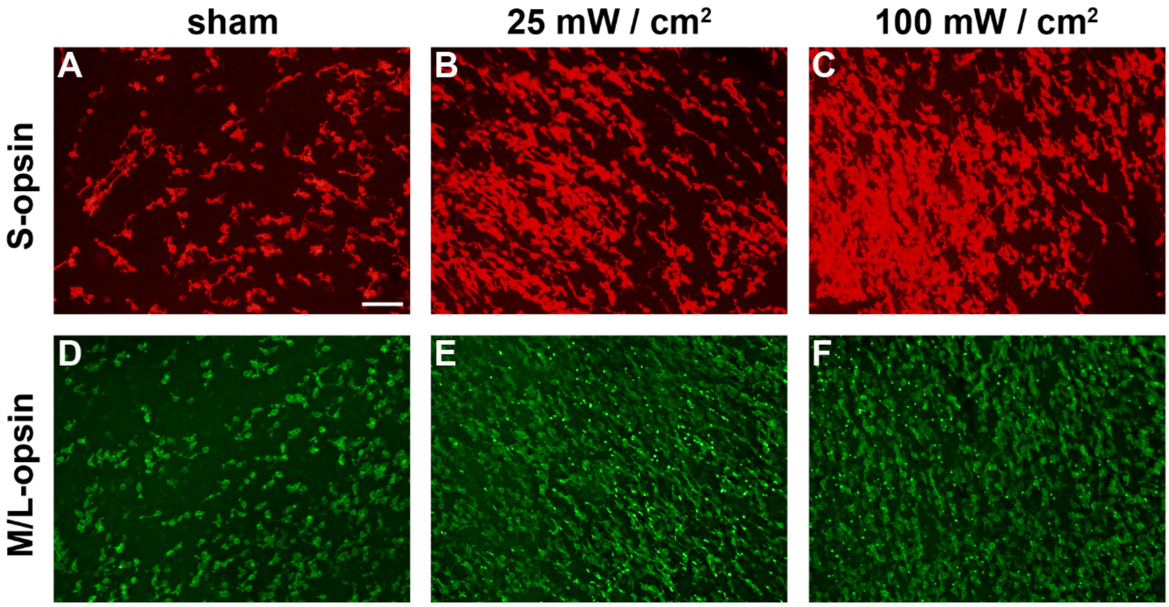


Supplementary Figure 2. Representative fluorescent photomicrographs of S-opsin$^{+}$ (A-C) and M/L-opsin$^{+}$ (D-F) immunoreactivities in *rd1* (P60) retinal wholemounts from the sham, 25 mW/cm$^{2}$ PBM and 100 mW/cm$^{2}$ PBM groups. Scale bar: 100 μm.

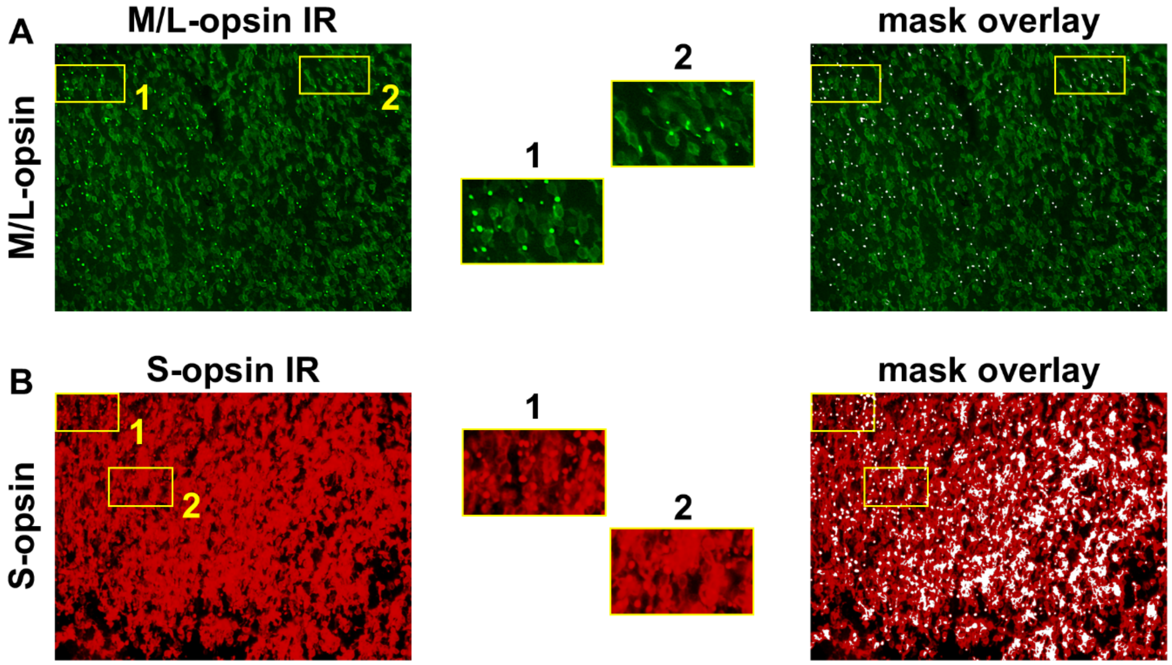


Supplementary Figure 3. Representative images of M/L-opsin$^{+}$ (A) and S-opsin$^{+}$ (B) immunoreactivities (IR) in retinal wholemounts from *rd1* mice. Insets 1 and 2 are magnified views of two regions from each photomicrograph. (A) For M/L-opsin$^{+}$, cell bodies stain lightly, but segments stain intensely (see insets). Thus, segments can be differentiated from cell bodies using image thresholding. The right panel shows the ML-opsin$^{+}$ IR overlaid with the mask derived from image thresholding (white represents areas to be quantified). It can be seen that the mask recapitulates the distribution of immunolabelled segments. (B) For S-opsin$^{+}$, cell bodies and segments both stain intensely (see insets). Thus, segments cannot be routinely differentiated from cell bodies using image thresholding, as illustrated by the S-opsin$^{+}$ IR overlaid with the mask derived from image thresholding. The mask does not recapitulate the distribution of immunolabelled segments. Thus, for quantification of outer segment survival, only ML-opsin$^{+}$ images were used, which represent both genuine ML-opsin$^{+}$ cones and dual cones, but not S-opsin$^{+}$ cones.
